# Brain Metastasis Screening Strategies in relation to Neurological Symptoms in Stage IV Lung Adenocarcinoma

**DOI:** 10.1101/2025.03.09.25323599

**Authors:** Sama I. Sayin, Ella A. Eklund, Moa Beischer, Torill Moe, Kevin X. Ali, Kerstin Gunnarsson, Moe Xylander, Lars Ny, Asgeir S. Jakola, Ida Häggström, Clotilde Wiel, Andreas Hallqvist, Volkan I. Sayin

## Abstract

**Background:** Brain metastases (BM) are a major clinical challenge in metastatic lung adenocarcinoma (LUAD), affecting up to 50% of patients during disease progression. Current guidelines do not mandate brain imaging for all metastatic lung cancer patients at diagnosis unless there are neurological symptoms present. However, real-world data on the predictive value of neurological symptoms for BM detection remain scarce.

**Methods:** This retrospective multicenter study analyzed all consecutive patients diagnosed with stage IV LUAD with molecular assessment in western Sweden from 2016-2021 (*n* = 912). We extracted data from patient charts, imaging referrals, radiology reports and the Swedish National Lung Cancer Registry to determine diagnostic brain imaging (DBI) frequency and modality, presence of neurological symptoms, BM detection rates, size, number, location and overall survival (OS).

**Results:** Among stage IV LUAD patients, 63% underwent DBI, and BM was detected in 23% of all patients (37% of those receiving DBI). Neurological symptoms prompted DBI in 63% of cases, yet 58% of these symptomatic patients had no BM on imaging. Conversely, 28% of asymptomatic patients who underwent DBI had BM. Patients with BM detected in the absence of neurological symptoms had smaller metastases. Neurological symptoms were associated with worse OS, irrespective of the presence of BM.

**Conclusion:** Neurological symptoms poorly predict BM in metastatic LUAD. Routine MRI-based brain imaging at diagnosis of metastatic disease, irrespective of neurological symptoms, may improve outcomes for this underserved patient population. These results provide real-world evidence supporting the need for reassessment of current BM screening recommendations.

## Introduction

Lung cancer remains the leading cause of cancer-related mortality worldwide, with advanced-stage patients facing a five-year survival rate below 10%, highlighting the urgent need for optimized clinical management. In Sweden, lung cancer is the fifth most common type of cancer but still accounts for the highest number of cancer-related deaths each year [1].

Lung adenocarcinoma (LUAD), the most common subtype of non-small cell lung cancer (NSCLC) and accounting for 50-60% of all lung cancer cases, poses significant clinical challenges, particularly when it metastasizes to the brain [2, 3]. Brain metastases (BM) occur in 25-30% of LUAD at diagnosis, and up to 50% of LUAD will develop BM during disease progression [4–6]. BM significantly worsens prognosis and often leads to debilitating neurological symptoms, impacting quality of life and requiring intervention of metastatic lesions in this organ to a larger extent compared to patients with metastases in other organ sites [7, 8].

While targeted therapy and immunotherapy are paving the way for novel approaches to treating BM, neurosurgical resection and stereotactic radiosurgery remain the most effective tools we have to date, and outcomes following both are adversely impacted by increasing tumor size and numbers [9]. Early detection of BM might improve outcomes, enabling timely multimodal treatment, particularly as the number of long-term BM survivors continues to rise together with improved treatment outcomes.

While it is standard practice to screen stage III NSCLC patients for BM before initiating curative treatment, stage IV patients without oncogenic driver mutations do not receive BM screening unless there are neurological symptoms. The European Society for Molecular Oncology (ESMO) guidelines for clinical management of metastatic lung cancer encourages brain imaging at diagnosis for all patients with metastatic disease, but specifies requirement of BM screening only for patients with neurological symptoms and signs [10]. However, real-world clinical data on whether the presence of neurological symptoms is a good indicator of the presence of BM in patients with extracranial metastatic disease are surprisingly lacking. Given the significant and increasing burden of BM in LUAD management, and the increasing role of multimodal treatment approaches, there is a pressing need for research into patterns of BM in relation to diagnostic brain imaging (DBI) and neurological symptoms to aid management decisions in the clinic.

In this retrospective multicenter study, we report real-world data on all consecutive patients diagnosed with metastatic LUAD with molecular assessment in western Sweden between 2016-2021. By combining data from patient charts, imaging referrals, radiology reports and the Swedish National Lung Cancer Registry, this study aims to provide a clearer understanding of the role of DBI in relation to neurological symptoms to inform future clinical management approaches in BM screening of stage IV LUAD.

## Materials and Methods

This study provides a real-world overview of how DBI was utilized in the clinical management of patients diagnosed with metastatic LUAD in western Sweden between 2016 and 2021. We conducted a multicenter retrospective study by combining data from the Swedish National Lung Cancer Registry with data from patient charts, imaging referrals and radiological reports for each patient. The Swedish national guidelines for lung cancer are in line with ESMO guidelines. By combining data about the frequency of DBI and whether the imaging was done due to neurological symptoms or not, we investigate whether symptoms is a good indicator for DBI. The Swedish healthcare system is primarily government-funded and provides universal access to all citizens. Therefore, all patients have equal access to diagnostic examinations and treatments.

### Patient Population

We included all consecutive lung cancer patients diagnosed with Stage IV LUAD and having molecular assessment performed between 2016–2021 in western Sweden (n = 912). One patient was excluded due to no DBI information being available, and one patient died between diagnostic sample collection and final diagnosis and is thus excluded from the overall survival (OS) analysis. Five patients included in the Swedish National Lung Cancer Registry did not have an available chart to investigate the presence of neurological symptoms and were therefore excluded from the analyses that used that information (Figure 1). Four patients did not have DBI modality reported, and three DBI reports of BM patients did not contain details of tumor size, location and diameter. Patient demographics (including age, gender, Eastern Cooperative Oncology Group (ECOG) performance status and smoking history) and outcome data were retrospectively collected. All patient charts were examined for whether a DBI was conducted or not. If DBI was done, the chart and imaging referral were examined to identify if it was done due to neurological symptoms being present or not. Neurological symptoms were defined as signs and symptoms that were deemed to be associated with BM according to clinical judgement, as specified in Supplementary Table 3. If the DBI detected brain metastasis, the number, location and largest diameter of metastases were collected from the DBI report. Approval from the Swedish Ethical Review Authority (Dnr 2019-04771 and 2021- 04987) was obtained prior to the commencement of the study. No informed consent was required due to all data being presented in a de-identified form according to the Swedish Ethical Review Authority.

**Figure 1.**
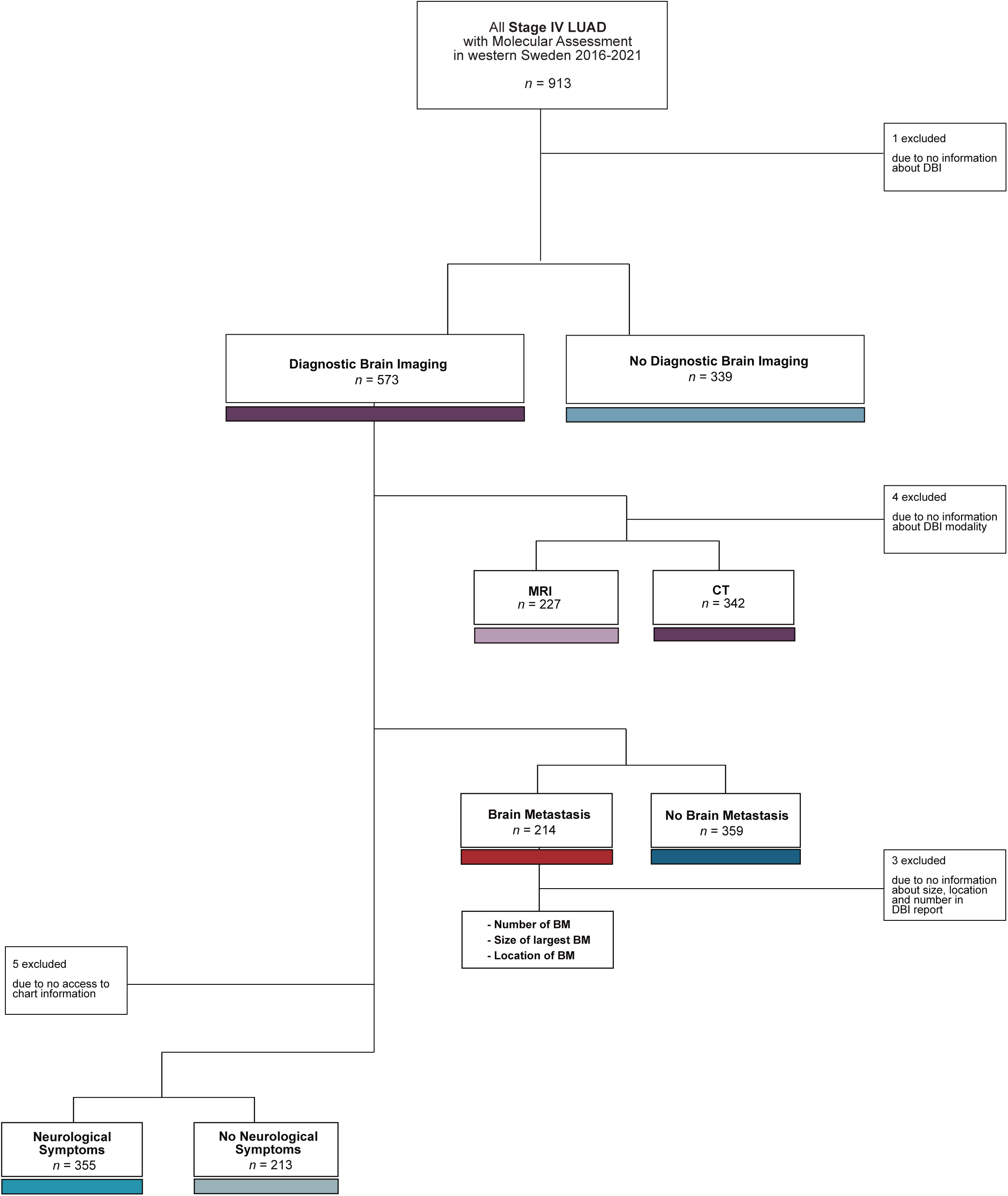
Flowchart showing patient selection for the study.

### Study Objectives

The primary outcome of this study was the frequency of DBI, neurological symptoms, presence of BM and OS, defined as the interval between the date of diagnostic sample collection of primary tumors and the date of death from any cause. Patients alive or lost to follow-up were censored at the cut-off date or last contact. Median follow-up time was 35 months (95% CI 31.1–38.9) and was estimated using the reverse Kaplan–Meier method. BM diagnosed within 8 weeks from date of diagnostic sample collection was considered as diagnosed at baseline. The data collection cut-off date was 2024-09-17.

### Statistical Analysis

Clinical characteristics were summarized using descriptive statistics and evaluated with univariate analysis in table form. Independent T-test and Pearson’s Chi-square test were used to identify differences in characteristics between groups. Survival was estimated using the Kaplan–Meier method. A log-rank test was used to assess significant differences in OS between groups. Statistical significance was set at *p* < 0.05, and no adjustments were made for multiple comparisons. Data analysis was conducted using IBM SPSS Statistics version 27 and R version 3.4.

## Results

### Patient characteristics

Among all stage IV LUAD patients, the majority (*n* = 573, 63%) received DBI (Table 1), and 23% (37% of group that received DBI) had BM (Supplementary Table 1 and Figure 2A). There were no significant differences in patient characteristics between the groups except that patients who had a DBI were slightly younger than those who did not (Table 1). Among all who received DBI, 60% received CT and 40% received MRI (Figure 2B). Importantly, there was no significant difference in OS between the groups that received DBI and those who did not (Figure 2C) and the DBI modality did not impact OS (Figure 2D).

**Figure 2.**
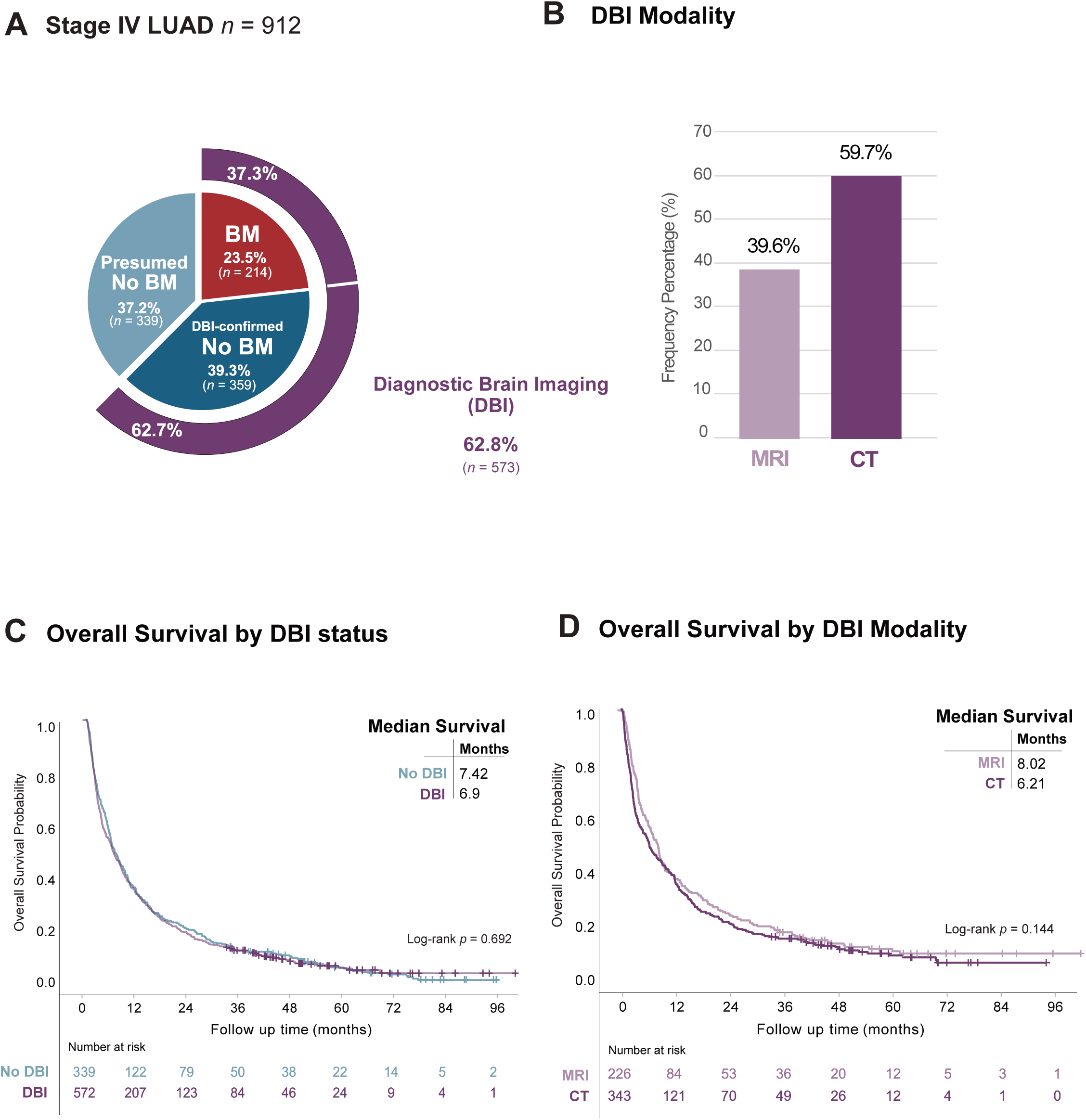
Diagnostic brain imaging (DBI) among patients with stage IV LUAD. **A)** Frequency distribution of DBI and brain metastasis (BM) in the study population (*n* = 912). **B)** Frequency of CT or MRI as modality of DBI. **C)** Kaplan-Meier estimates comparing OS stratified by presence (purple) or absence (blue) of DBI. **D)** Kaplan-Meier estimates comparing OS stratified by CT (dark purple) or MRI (light purple) as modality of DBI.

**Table 1.** Patient characteristics of entire study population - stratified by presence of diagnostic brain imaging (DBI).

| Patient characteristics<br>(All Stage IV LUAD) | Total | No DBI | DBI | p-value |
| --- | --- | --- | --- | --- |
|  | n (%) | n (%) | n (%) |  |
| All subjects | 912 (100) | 339 (37.2) | 573 (62.8) |  |
| Age in years mean (range) | 71 (26-94) | 72 (31-94) | 70 (26-91) | < 0.001 |
| Sex |  |  |  | 0.421 |
| Male | 404 (44.3) | 156 (46.0) | 248 (43.3) |  |
| Female | 508 (55.7) | 183 (54.4) | 325 (56.7) |  |
| Smoking history |  |  |  | 0.158 |
| Current smoker | 280 (30.7) | 92 (27.1) | 188 (32.8) |  |
| Former smoker | 438 (48.0) | 169 (49.9) | 269 (46.9) |  |
| Never smoker | 191 (21.0) | 78 (23.0) | 113 (19.7) |  |
| Missing | 3 (0.3) | 0 | 3 (0.5) |  |
| Performance status |  |  |  | 0.311 |
| ECOG 0 | 101 (11.1) | 38 (11.2) | 63 (11.0) |  |
| ECOG 1 | 341 (37.4) | 117 (34.5) | 224 (39.1) |  |
| ECOG 2 | 244 (26.8) | 92 (27.1) | 152 (26.5) |  |
| ECOG 3 | 144 (15.8) | 63 (18.6) | 81 (14.1) |  |
| ECOG 4 | 34 (3.7) | 10 (2.9) | 24 (4.2) |  |
| Missing | 48 (5.3) | 19 (5.6) | 29 (5.1) |  |
| Histology |  |  |  |  |
| Adenocarcinoma | 912 (100) | 339 (100) | 573 (100) |  |
| Mutation status |  |  |  |  |
| None known | 340 (37.3) | 129 (38.1) | 211 (36.8) | 0.711 |
| KRAS G12C | 140 (15.4) | 41 (12.1) | 99 (18.0) | 0.036 |
| KRAS Other | 187 (20.5) | 84 (24.8) | 103 (18.0) | 0.014 |
| EGFR | 146 (16.0) | 49 (14.5) | 97 (16.9) | 0.325 |
| ALK | 34 (3.7) | 10 (2.9) | 24 (4.2) | 0.340 |
| BRAF | 43 (4.7) | 17 (5.0) | 26 (4.5) | 0.742 |
| ROS1 | 23 (2.4) | 7 (2.1) | 15 (2.6) | 0.599 |
| HRAS Q61R | 12 (1.3) | 5 (1.5) | 7 (1.2) | 0.746 |
| MET | 8 (0.9) | 5 (1.5) | 3 (0.5) | 0.136 |
| PIK3CA | 4 (0.4) | 3 (0.9) | 1 (0.2) | 0.117 |
| RET | 3 (0.3) | 0 | 3 (0.5) | 0.182 |
| JAK2 | 1 (0.1) | 1 (0.3) | 0 | 0.193 |
| PD-L1 grade (%) |  |  |  | 0.354 |
| 0 | 470 (51.5) | 183 (54.0) | 287 (50.1) |  |
| ≥ 1 | 140 (15.4) | 55 (16.2) | 85 (14.8) |  |
| ≥ 20 | 85 (9.3) | 26 (7.7) | 59 (10.3) |  |
| ≥ 50 | 271 (23.8) | 75 (22.1) | 142 (24.8) |  |
| At last follow up |  |  |  | 0.163 |
| Alive | 83 (9.1) | 25 (7.4) | 58 (10.1) |  |
| Deceased | 829 (90.9) | 314 (92.6) | 515 (89.9) |  |
| Survival |  |  |  |  |
| Median survival (months) | 7 | 7 | 7 | 0.711 |

### DBI findings in BM patients

The majority (70%) of BM was diagnosed with MRI, while around a third (29.9%) received CT (Figure 3A). At diagnosis, 32% of BM patients had 1 BM, with fewer patients presenting with increasing numbers of BM up to 9 tumors. The proportion of patients with more than 10 BM at diagnosis was 9% (Figure 3B). The most common location of BM was supratentorial (53%) followed by having both supra- and infratentorial BM (36%) and only infratentorial BM was rare (9%) (Figure 3C). When looking at the size of the largest BM at diagnosis, 34% of all BM on DBI were 11-20 mm, and 60% BM were less than 30 mm in diameter (Figure 3D).

**Figure 3.**
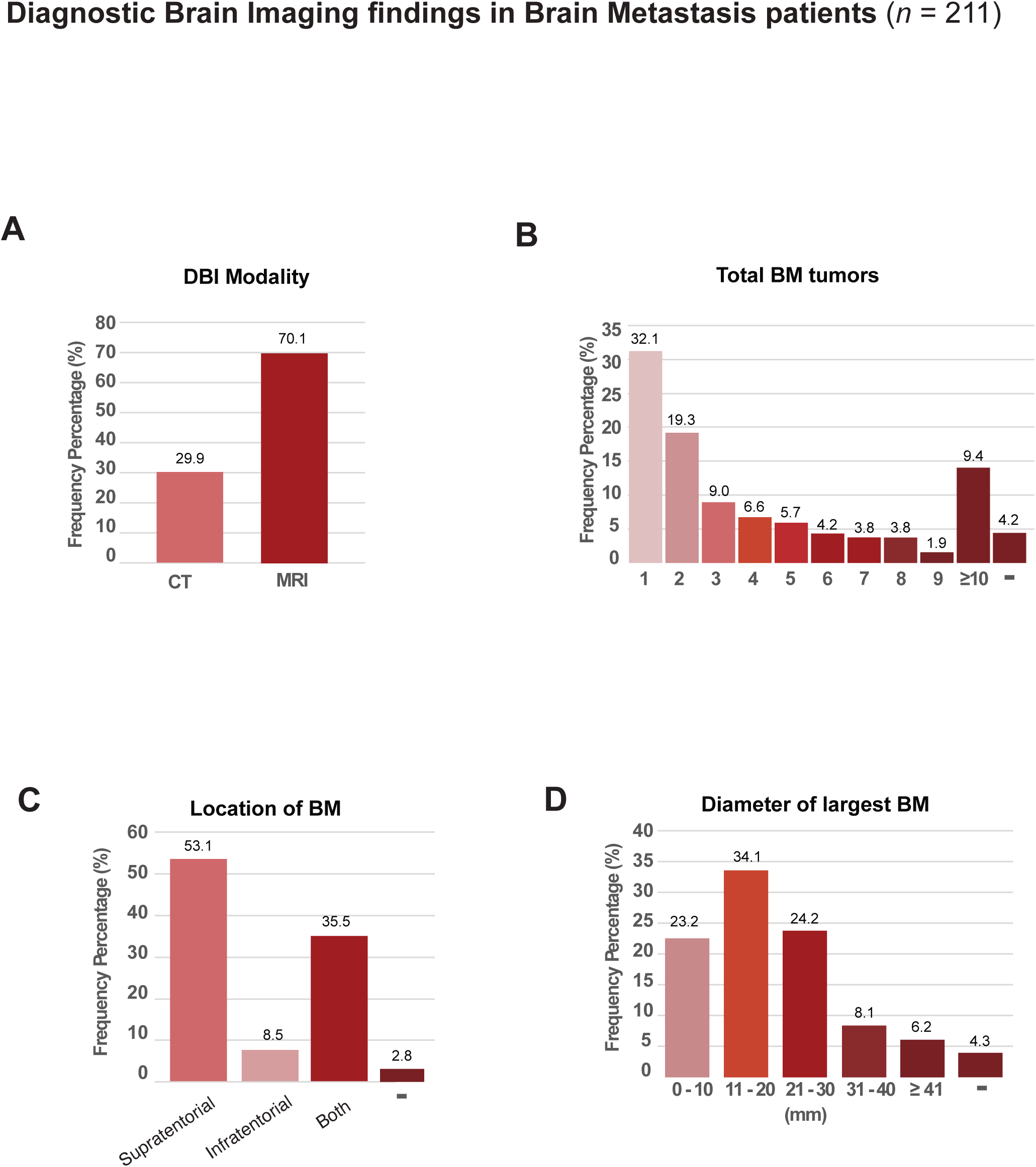
DBI findings in stage IV LUAD patients with BM. (-) Not specified in charts or DBI report for **B)** Number, **C)** Location, or **D)** Numerical size measurement.

### Neurological symptoms among stage IV LUAD with DBI

Next, we investigated whether the DBI among stage IV LUAD patients were received due to the presence of neurological symptoms. We found that the majority (63%) of DBI referrals were due to neurological symptoms (Table 2 and Figure 4A). The group with symptoms were older (*p* < 0.001) and had a worse performance status with the majority being ECOG ≥2 (*p* = 0.005) (Table 2). Additionally, patients with neurological symptoms had significantly worse OS with a median of 5 compared to 12 months for those without symptoms (*p* <0.0001) (Figure 4B).

**Figure 4.**
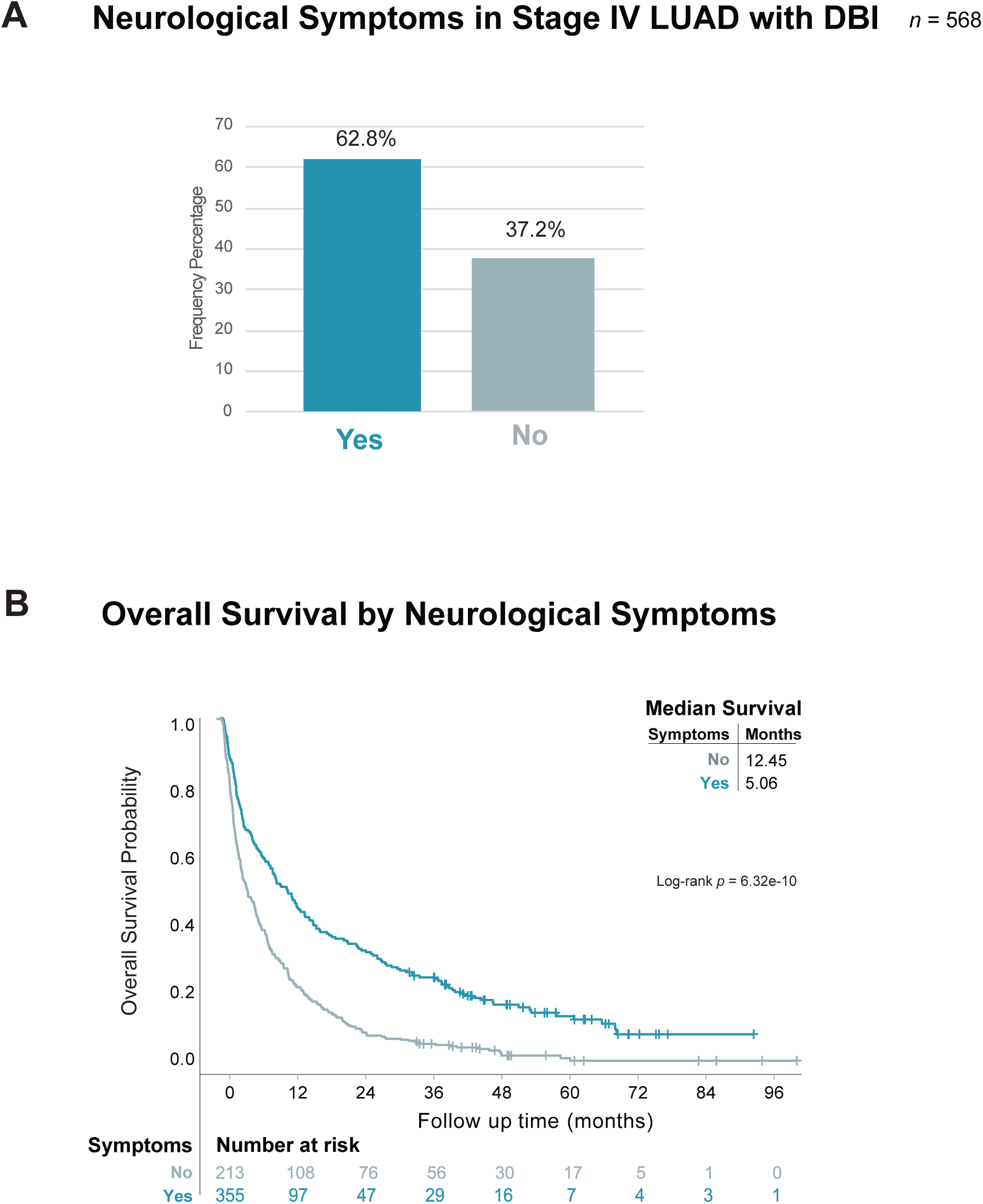
Neurological symptoms in stage IV LUAD with DBI. **A)** Frequency of presence (Yes) or absence (No) of neurological symptoms in the study population (*n* = 568). **B)** Kaplan-Meier estimates comparing OS stratified by presence (Yes) or absence (No) of neurological symptoms.

**Table 2.**
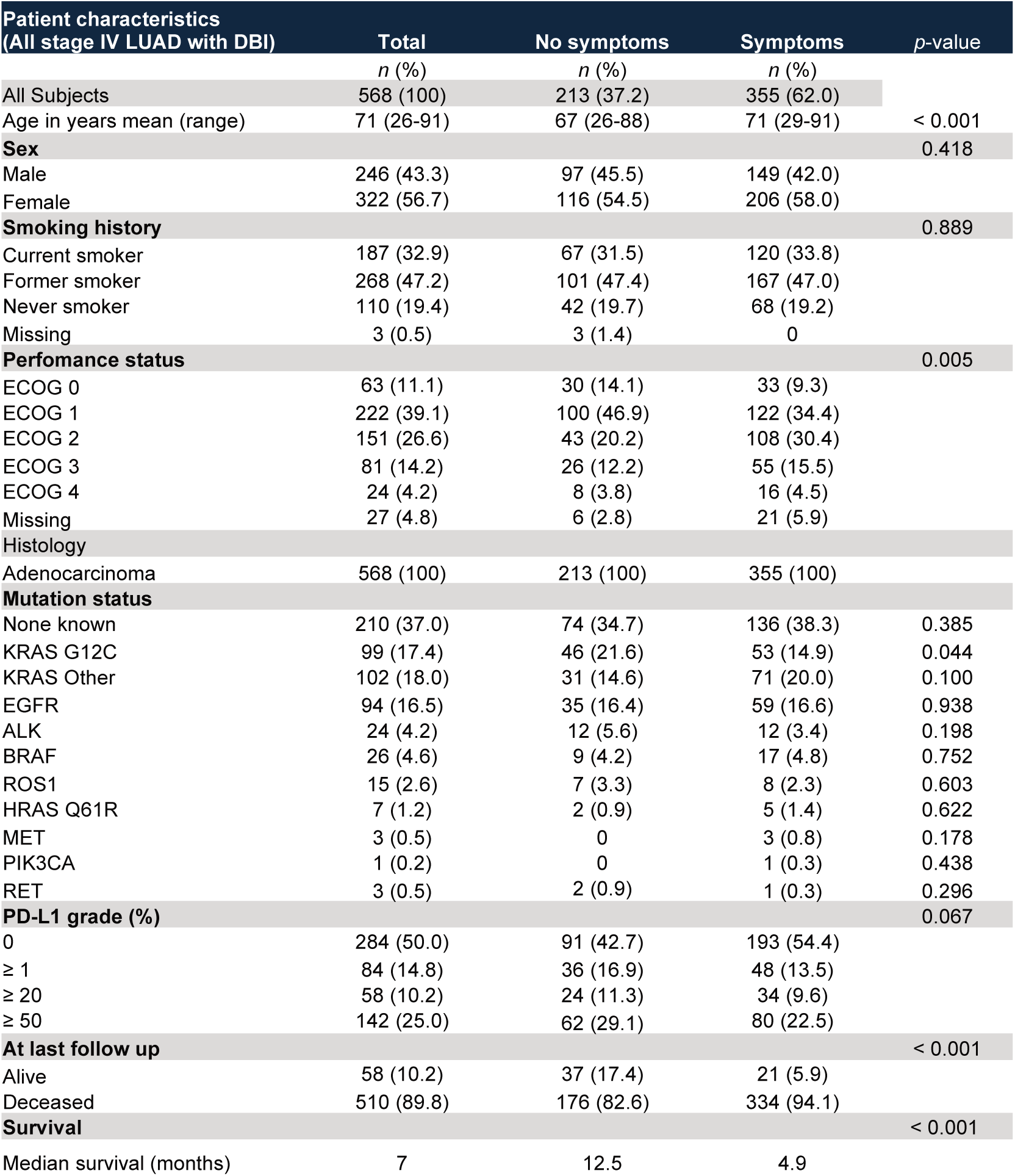
Patient characteristics of all stage IV LUAD patients with DBI - stratified by presence or absence of neurological symptoms.

### Neurological symptoms in relation to BM detection at diagnosis in LUAD

Next, to probe the value of neurological symptoms as an indicator for diagnostic BM screening, we studied DBI findings in the subgroups with and without symptoms. In the absence of neurological symptoms, 70% received CT and 30% received MRI, while higher proportion (46%) received an MRI when neurological symptoms were present (Figure 5A).

**Figure 5.**
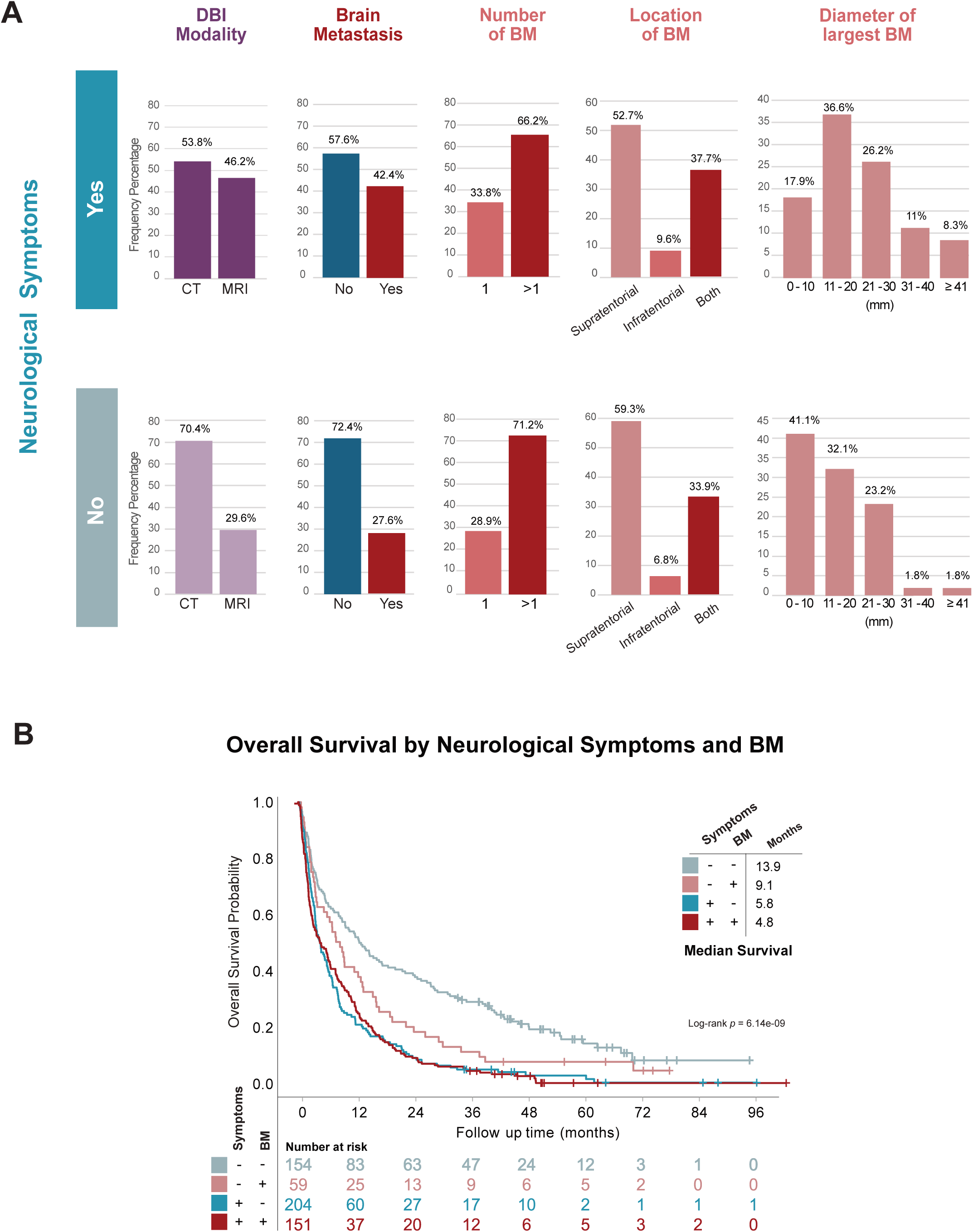
Neurological symptoms in relation to BM in stage IV LUAD with DBI. **A)** Frequency distribution of DBI findings in stage IV LUAD in relation to Presence (Yes, top panels) or absence (No, bottom panels) of neurological symptoms: imaging modality (purple), presence of BM (red or blue), and DBI findings in BM patients (shades of red). **B)** Kaplan-Meier estimates comparing OS stratified by presence or absence of neurological symptoms and of BM.

Importantly, 58% of all stage IV with neurological symptoms at diagnosis had no BM on DBI, while 28% of those who received DBI even in the absence of any neurological symptoms at diagnosis had BM (Figure 5B).

Distribution of number or location of BM were similar between groups regardless of neurological symptoms. However, when looking at size of the largest BM tumor, we found that while only 18% of BM tumors under 10 mm were detected when DBI was performed in the presence of neurological symptoms, up to 41% BM were detected with tumors less than 10 mm in size when DBI was performed even though no neurological symptoms were present (Figure 5A).

### Neurological symptoms impact OS independent of BM

Further we investigated the impact of the presence of neurological symptoms in relation to whether a BM was detected with DBI. As expected, patients without symptoms and no BM had a longer median OS of 14 months (Figure 5B). However, the patients presenting symptoms without BM had almost as short median OS as the patients with symptoms and BM with a median of 5.8 vs 4.8 months (*p =* 1.00) (Figure 5B and Supplementary Table 2). The patients without neurological symptoms and BM had a numerically better OS than the patients with symptoms and BM with a median of 9.1 months vs 4.8 months (*p* = 0.253) (Figure 5B and Supplementary Table 2).

## Discussion

Current ESMO guidelines recommend brain imaging at diagnosis for all patients with metastatic lung cancer and mandate it for those with neurological symptoms or signs [10]. This real-world multi-center study investigated the patterns of clinical adherence to this recommendation. About a third of the patients receiving DBI did not present any neurological symptoms. However, a relatively large proportion of asymptomatic patients had BM present. Compared with those with neurological symptoms, smaller BMs were detected when DBI was performed in the absence of neurological symptoms, opening avenues for more treatment possibilities in this subgroup through early detection.

To our knowledge our study design is unique as there have not been any directly comparable studies found in literature. While most studies regarding BM focus on detecting risk factors for BM or the prognostic impact of BM in lung cancer, studies evaluating real-world impact of administering DBI dependent on neurological symptoms are lacking. In line with our results, Ohhara et. al. [11] reported longer OS in the absence of symptoms with BM compared to in the presence of symptoms and BM. Dubbé-Pelletier et al. also studied stage IV patients without symptoms and found 30% with BM at diagnosis, comparable to our results [12]. Importantly, previous studies investigating presence of BM in relation to DBI have excluded patients with neurological symptoms, and we show here that presence of symptoms is not a reliable indicator of the presence of BM (58% of patients without symptoms had no BM on DBI).

Importantly, stage IV patients with neurological symptoms had worse OS regardless of the presence of BM. This group was systematically different from those without symptoms, with increased age and worse PS. In the subgroup of patients with symptoms without BM that still had worse survival, comorbidities such as leptomeningeal disease and treatment-associated toxicity may be driving outcomes. Also, there are probably several BM patients hidden in this group, for example, those with CT only. This patient group should be further characterized in future studies to investigate BM-independent mechanisms of neurological symptoms associated with worse prognosis in stage IV LUAD.

Our results also show a preference for CT in the absence of neurological symptoms, although MRI has been shown to be more effective and should be prioritized according to guidelines [13]. In addition, Waizman et al found that asymptomatic patients with good PS who undergo MRI have improved survival [14]. This indicates that there is a risk that, in patients only undergoing CT, BM could have been missed that perhaps would have been detected on MRI.

While EGFR and ALK-mutated stage IV NSCLC now receive routine DBI, our dataset still showed comparable number of patients with these mutations with the group that did not receive DBI. Additionally, there was no enrichment of EGFR or ALK even in the patients receiving a DBI without symptoms. This is likely due to guidelines requiring DBI for all EGFR and ALK-mutated stage IV were updated during the study period.

Given our results, new treatment strategies like SRS of several BMs, newer systemic therapies with improved blood-brain barrier penetrance, and as screening is already routine for liver and adrenal metastases in this group, routine brain imaging at diagnosis with MRI, regardless of the presence of neurological symptoms, is likely warranted for all patients with metastatic lung cancer.

### Limitations

The findings and conclusions drawn from this study are limited by its retrospective nature. While prospective and larger studies may be required to substantiate these findings with better control of confounders, we know that proper staging including brain imaging is beneficial for stage III patients and is already conducted for stage IV patients with oncogenic drivers. Additionally, the stratification of groups according to the presence of neurological symptoms is also limited by its definition, and confounders such as comorbidities and treatment modalities have not been compensated for.

## Conclusion

Presence of neurological symptoms is a bad indicator for BM screening at diagnosis of stage IV LUAD. Early detection of BM through routine brain imaging in all stage IV LUAD patients irrespective of neurological symptoms can facilitate prompt and appropriate treatment, potentially leading to improved outcomes for this historically underserved patient population.

## Ethical Statement

Approval from the Swedish Ethical Review Authority (Dnr 2019-04771 and 2021-04987) was obtained prior to the commencement of the study. No informed consent was required due to all data being presented in a de-identified form.

## Consent for publication

Not applicable. Patient consent statements were not required due to the retrospective nature of this study.

## CRediT Author contributions

**SIS**: Conceptualization; Formal analysis; Investigation; Methodology; Supervision; Validation; Visualization; Writing - original draft; and Writing - review & editing

**EAE**: Conceptualization; Formal analysis; Investigation; Methodology; Supervision; Validation; Visualization; Writing - original draft; and Writing - review & editing

**MB** and **TM**: Data curation; Formal analysis

**KG**: Methodology; Writing - review & editing

**KXA**: Formal analysis; Methodology; Visualization

**MX**: Visualization; Writing - review & editing

**LN, ASJ:** Conceptualization; Writing - review & editing

**IH, CW:** Conceptualization; Funding acquisition; Project administration; Writing - review & editing

**AH, VIS:** Conceptualization; Project administration; Supervision; Funding acquisition; Writing - review & editing

## Data availability

The datasets used and/or analyzed during the current study is available from the corresponding author on reasonable request.

## Declaration of generative AI and AI-assisted technologies in the writing process

During the preparation of this work the authors used ChatGPT4o to proof text and improve language for clarity. After using this tool, the authors reviewed and edited the content as needed and take full responsibility for the content of the publication.

---

**Supplementary Table 1.** Patient characteristics of all stage IV LUAD patients with DBI - stratified by presence or absence of brain metastasis (BM). p-values are from Pearson’s Xi.

| Patient characteristics<br>(All stage IV LUAD with DBI) | Total | No BM | BM | p-value |
| --- | --- | --- | --- | --- |
|  | n (%) | n (%) | n (%) |  |
| All subjects | 573 (62.8) | 359 (62.7) | 214 (37.3) |  |
| Age in years mean (range) | 71 (26-91) | 70 (26-91) | 69 (34-87) | 0.041 |
| **Sex** |  |  |  | 0.043 |
| Male | 248 (43.3) | 167 (46.5) | 81 (37.9) |  |
| Female | 325 (56.7) | 192 (53.5) | 133 (62.1) |  |
| **Smoking history** |  |  |  | 0.461 |
| Current smoker | 188 (32.8) | 111 (30.9) | 77 (36.0) |  |
| Former smoker | 269 (46.9) | 173 (48.2) | 96 (44.9) |  |
| Never smoker | 113 (19.7) | 73 (20.3) | 40 (18.7) |  |
| Missing | 3 (0.5) | 2 (0.6) | 1 (0.5) |  |
| **Performance status** |  |  |  | 0.469 |
| ECOG 0 | 63 (11.0) | 36 (10.0) | 27 (12.6) |  |
| ECOG 1 | 224 (39.1) | 141 (39.3) | 83 (38.8) |  |
| ECOG 2 | 152 (26.5) | 91 (25.3) | 61 (28.5) |  |
| ECOG 3 | 81 (14.1) | 57 (15.9) | 24 (11.2) |  |
| ECOG 4 | 24 (4.2) | 16 (4.5) | 8 (3.7) |  |
| Missing | 29 (5.1) | 18 (5.0) | 11 (5.1) |  |
| **Histology** |  |  |  |  |
| Adenocarcinoma | 573 (100) | 359 (100) | 214 (100) |  |
| **Mutation status** |  |  |  |  |
| None known | 211 (36.8) | 133 (37.0) | 78 (36.4) | 0.886 |
| KRAS G12C | 99 (18.0) | 65 (18.1) | 34 (15.9) | 0.497 |
| KRAS Other | 103 (18.0) | 69 (19.2) | 34 (15.9) | 0.315 |
| EGFR | 97 (16.9) | 48 (13.4) | 49 (22.9) | 0.003 |
| ALK | 24 (4.2) | 15 (4.2) | 9 (4.2) | 0.987 |
| BRAF | 26 (4.5) | 21 (5.8) | 5 (2.3) | 0.051 |
| ROS1 | 15 (2.6) | 14 (3.9) | 1 (0.5) | 0.013 |
| HRAS Q61R | 7 (1.2) | 3 (0.8) | 4 (1.9) | 0.276 |
| MET | 3 (0.5) | 1 (0.3) | 2 (0.9) | 0.293 |
| PIK3CA | 1 (0.2) | 0 | 1 (0.5) | 0.195 |
| RET | 3 (0.5) | 3 (0.8) | 0 | 0.180 |
| **PD-L1 grade (%)** |  |  |  | 0.341 |
| 0 | 287 (50.1) | 170 (47.4) | 117 (54.7) |  |
| ≥ 1 | 85 (14.8) | 59 (16.4) | 26 (12.1) |  |
| ≥ 20 | 59 (10.3) | 39 (10.9) | 20 (9.3) |  |
| ≥ 50 | 142 (24.8) | 91 (25.3) | 51 (23.8) |  |
| **At last follow up** |  |  |  | 0.105 |
| Alive | 58 (10.1) | 42 (11.7) | 16 (7.5) |  |
| Deceased | 515 (89.9) | 317 (88.3) | 198 (92.5) |  |
| **Survival** |  |  |  | 0.038 |
| Median survival (months) | 7 | 8 | 6 |  |

**Supplementary Table 2.** Pairwise Log-Rank Test p-values for comparison of OS groups in Figure 5B.

| Comparison groups | p-value |
| --- | --- |
| · No Symptoms & BM vs No Symptoms & No BM | 0.262 |
| · Symptoms & No BM vs No Symptoms & No BM | 2.36E-08 |
| · Symptoms & BM vs No Symptoms & No BM | 8.45E-07 |
| · Symptoms & No BM vs No Symptoms & BM | 0.198 |
| · Symptoms & BM vs No Symptoms & BM | 0.253 |
| · Symptoms & BM vs Symptoms & No BM | 1 |

**Supplementary Table 3.** List of chart details considered to indicate Neurological Symptoms.

| List of chart details defined as **Neurological Symptoms**: |
| --- |
| • Headaches.<br>• Seizures. Such as episodes of numbness, tingling, uncontrollable arm and leg movements, difficulty speaking, strange smells or sensations, staring and unresponsive episodes or convulsions.<br>• Vertigo<br>• Loss of balance or coordination.<br>• Changes in mental function, mood or personality.<br>• Weakness or numbness in the arms or legs.<br>• Changes in the senses: Impaired ability to hear, smell or see. This can include double vision or blurred vision.<br>• Difficulty speaking or understanding language.<br>• Changes in the sense of touch: Impaired ability to feel heat, cold, pressure, a light touch or sharp objects may change. |

